# Is it Just About Physical Health? An Online Cross-Sectional Study Exploring the Psychological Distress Among University Students in Jordan in the midst of COVID-19 Pandemic

**DOI:** 10.1101/2020.05.14.20102343

**Authors:** Ala’a B. Al-Tammemi, Amal Akour, Laith Alfalah

**Author notes:** **Correspondence:** Ala’a B. Al-TammemiAddress 1:Historiegränd 90734, Umeå, Sweden; Address 2:Laktanya utca 4028, Debrecen, Hungary.

## Abstract

**Background:** Since the spread of COVID-19 on a global scale, most of efforts at national and international levels were directed to mitigate the spread of the disease and its physical harm, paying less attention to the psychological impacts of COVID-19 on global mental health especially at early stages of the pandemic.

**Objectives:** This study aimed to assess and explore (i) The levels of psychological distress and its correlates (ii) Motivation for distance learning (iii) Coping activities and pandemic related concerns, among university students in Jordan in the midst of COVID-19 pandemic

**Methods:** A cross-sectional study was conducted using an online self-administered questionnaire. The measure of psychological distress was obtained using the 10-item Kessler Psychological Distress Scale,while other questions have explored our study’s second and third aims.

**Results:** A total of 381 completed questionnaires were included in the analysis. Female participants slightly predominated the sample (n=199, 52.2%). The respondents aged 18-38 years (mean 22.6 years, SD: 3.16). Concerning distress severity, most of respondents were regarded as having severe psychological distress (n=265, 69.5%). 209 students (54.9%) reported that they had no motivation for distance learning. Ordinal logistic regression revealed a significant correlation between distress severity and many predictors. Among the predictors that were found to act as a protective factors against higher levels of distress included older age (aOR=0.64, P=0.022; 95% CI: 0.44 - 0.94), and having a strong motivation for distance learning (aOR=0.10, P=0.048 ; 95% CI: 0.01 - 0.96). In contrary, being a current smoker (aOR=1.99, P=0.049 ; 95% CI: 1.10 - 3.39), and having no motivation for distance learning (aOR=2.49, P=0.007; 95% CI: 1.29 - 4.80) acted as risk factors for having higher levels of psychological distress among the students. The most common coping activity reported was spending more time on social media platforms (n=269, 70.6%), and 209 students (54.9%) reported distance learning was their most distressing concern.

**Conclusion:** The COVID-19 pandemic and related control measures could impact the mental health of individuals, including students. We recommend a nationwide psychological support program to be incorporated into Jordan’s preparedness plan and response strategy in combating the COVID-19 pandemic.

## 1 INTRODUCTION

COVID-19 is a highly transmissible respiratory disease caused by a new type of human coronaviruses; SARS-CoV-2 (Al-Tammemi, 2020). Since its discovery in late December 2019, the disease has spread widely across many countries and territories on a global scale. As of September 20,2020 more than 30 million confirmed cases, and over nine hundred thousand confirmed deaths across 216 countries and territories were attributed to the COVID-19 (World Health Organization, 2020).

Epidemics and outbreaks can pose profound impacts on physical health, mental health as well as the global economy resulting in disruptions of humans’ daily life (Chakraborty and Maity, 2020). The containment measures that were adopted by many countries worldwide in combating the COVID-19 such as quarantine, countries’ lockdown, travel restrictions, physical distancing, social isolation as well as local restrictions on individuals’ mobility, can lead to a significant burden on mental health causing emotional and behavioral changes (SAMHSA, 2014; Brooks et al., 2020; Cao et al., 2020; Center for the Study of Traumatic Stress, 2020; Holmes et al., 2020).

In addition, the psychological impacts of outbreaks are considered a threat not only on individuals with pre-existing psychiatric illness but also on those who are free of any psychiatric condition (Ho et al., 2020). The fear of an epidemic can afflict individuals irrespective of their gender, age, race, or socioeconomic status. Anxiety, insomnia, anger, loneliness, fear, shame, helplessness, blame, guilt, and stigma were all found to be present during infectious diseases’ outbreaks (Ho et al., 2020; Ornell et al., 2020). Different psychiatric conditions, including depression, panic attacks, Post Traumatic Stress Disorder, and even suicidality, were also reported to be associated with outbreaks, especially in younger age groups (Ho et al., 2020).

In epidemics, certain groups in the society such as older people, children, health care workers, infected patients, patients with pre-existing psychiatric conditions and students are at a greater risk of suffering from a significant degree of psychological pressure and stress compared to other individuals (Ho et al., 2020). It is essential to gather information about the impacts of the COVID-19 pandemic on the mental health of the general population and specific vulnerable groups, and this will help in developing appropriate interventions that would mitigate such pandemic’s adverse effects (Holmes et al., 2020). Since the beginning of the COVID-19 pandemic, most of the global efforts act on the biological and physical aspects of the pandemic in order to limit its spread within the communities. However, much less attention was paid to the mental health risks of the COVID-19 pandemic especially at early stages of the pandemic.

Jordan is amongst the countries that have been struck by the COVID-19 pandemic, and in response to that, many preventive and control strategies were enforced by the government to retard the viral spread in the country. One of Jordan’s public health responses during early stages of the pandemic was declaring the closure of all schools and higher academic institutions with shifting to online remote learning since the middle of March 2020 (Al-Tammemi, 2020; Jordanian Ministry of Health, 2020; Prime Ministry of Jordan, 2020). The COVID-19 pandemic alongwith the disruptions that happened in various sectors including the academic sector has forced the students to live in a new experience at both academic and personal levels. Consequently and in light of limited literatures that assessed mental health status of university students in Jordan, our present study aimed at (i) Exploring the level of psychological distress and its correlates amongst university students during the COVID-19 pandemic (ii) Evaluating the students’ motivation for distance learning and, (iii) Exploring coping activities and major pandemic related concerns from students’ perspective.

## 2 MATERIALS AND METHODS

### 2.1 Study Design and Participants

A cross-sectional study was conducted in May 2020, using an online self-administered questionnaire of closed-ended questions. The participants in our study were recruited through social media platforms employing a convenience sampling strategy. The questionnaire was distributed across seven randomly selected Facebook groups of university students in Jordan and academic groups on WhatsApp messenger for a duration of one day. These social media groups were created by students as a tool for general and academic communication within the students’ community and involved students who are currently enrolled in different study programs and levels at various academic institutions in Jordan. The students who were available and voluntarily willing to be involved in the study could open a link to get an information letter about the study, eligibility criteria, and informed consent as a prerequisite to proceed in participation.Considering the nature of the web-based Google form surveys, the students were instructed to fill out the questionnaire with probity after fullfilling the eligibility criteria, consenting on voluantry participation and filling it only once. We did not provide any form of compensation to the participants upon their involevement in our study.

We decided to carry out this study using an internet-based survey due to the current pandemic crisis and the national strict measures on the face to face communication coupled with the closure of all academic institutions in Jordan at the time of data collection In addition, using the internet and social media for the recruitment and sampling procedures in this study has shown to be an effective and time-efficient method to reach inaccessible potential participants from different Jordanian regions by eliminating any geographical boundaries. A recent systematic review of 109 published articles that aimed at evaluating the use of social media such as Facebook for recruitment of research participants in various psychological and medical studies came into evidence, which supported the effectiveness and efficiency of this strategy (Thornton et al., 2016).

For a student to be able to participate in this study, all the following eligibility criteria were implemented:

1. Age ≥ 18 years
2. Residing in Jordan during the pandemic crisis
3. Active enrollment in an undergraduate or postgraduate study at a Jordanian University.

### 2.2 Instruments and Measures

The online questionnaire was created using *Google Forms* provided by Google (tm) and was constructed in modern standard Arabic. The questionnaire consisted of three sections, with a total of 24 questions. The first section comprised of seven questions about sociodemographic information including age, gender, region of residence, study level, type of academic institution, marital status, and smoking status alongwith two questions about any history of pre-existing psychiatric conditions and related medication use.

The second section included an Arabic version of the 10-item Kessler Psychological Distress Scale (K10). This Arabic version was translated from the original English version by a team of linguistic experts from multiple Arab countries (Egypt, Libya, Lebanon, and Tunisia) in addition to Arab experts in Psychology in the United States. The Arabic version is provided by Harvard Medical School on the webpage of the National Comorbidity Survey (National Comorbidity Survey, 2013).

The 10-item Kessler Psychological Distress Scale (K10) is an internationally validated tool for simple and rapid assessment/screening of non-specific psychological distress in which 10 questions with 5-point Likert scale responses are present (Andrews and Slade, 2001; Kessler et al., 2002; Fassaert et al., 2009; Easton et al., 2017). On a sample of Arabs, the Arabic version of the 10-item Kessler Psychological Distress Scale (K10) has shown satisfactory psychometric properties with high internal consistency and reliability (Cronbach’s α = 0.88) (Easton et al., 2017).

The questions of the 10-item Kessler Psychological Distress Scale (K10) are:

**Question 1 (Q1)**. “During the last 30 days, about how often did you feel tired out for no good reason?”

**Question 2 (Q2)**. “During the last 30 days, about how often did you feel nervous?”

**Question 3 (Q3)**. “During the last 30 days, about how often did you feel so nervous that nothing could calm you down?”

**Question 4 (Q4)**. “During the last 30 days, about how often did you feel hopeless?”

**Question 5 (Q5)**. “During the last 30 days, about how often did you feel restless or fidgety?”

**Question 6 (Q6)**. “During the last 30 days, about how often did you feel so restless you could not sit still?”

**Question 7 (Q7)**. “During the last 30 days, about how often did you feel depressed?”

**Question 8 (Q8)**. “During the last 30 days, about how often did you feel that everything was an effort?”

**Question 9 (Q9)**. “During the last 30 days, about how often did you feel so sad that nothing could cheer you up?”

**Question 10 (Q10)**. “During the last 30 days, about how often did you feel worthless?”

The response choices with their correspondence score weights are *None of the time* (1 point), *A little of the time* (2 points), *Some of the time* (3 points), *Most of the time* (4 points), and *All the of time* (5 points). With having 10 questions and five weighted responses as previously described, the total minimum and maximum scores for the Kessler distress scale (K10) are 10 and 50, respectively. As per the scale’s guide, **Q3** and **Q6** were not asked in our study and were automatically scored as one point if the preceding questions **Q2** and **Q5** were answered as *None of the time*.

The severity of psychological distress was then categorized into four groups as the following based on the total K10 distress score for each participant: 10-19 = no psychological distress, 20-24 = mild psychological distress, 25-29 = moderate psychological distress, and 30-50 = severe psychological distress (Andrews and Slade, 2001).

The third section of the questionnaire included five questions about the following topics: one question about *coping activities during COVID-19 pandemic and the nationwide curfew in Jordan*.This question included a list of 13 activities from which the students were able to choose all that applies to their situation and to add any activity that was not listed among the choices using the option *“others, please specifiy”*. Most of the listed acitivities were suggested by the authors and few others were adapted from another resource (USCF, 2020).Amongst these activities were spending more time on social networking platforms, talking to friends, watching television, more engagement with family, listening to music, practicing sports at home, studying and preparing for exams, increase smoking, reading Books / novels, meditation, herbal drinks, practicing Yoga, talking to a psychological counselor and others. Two questions about *the use of medications to cope with COVID-19 related distress*,in which one of the questions was with yes/no response to know whether the student used a medicinal drug to cope with pandemic distress or not and if yes, to report the frequency of usage, while the other question included different classes of medications with examples on most common trade names in each class and the students could add any medication that was not listed using the option *“others, please specifiy”*. Additionally, one question about *students’ motivation for online distance learning*, using a single-answer item with responses as no motivation, low motivation, moderate motivation, and strong motivation, and lastly, a question about *major pandemic related concerns as perceived by the students*. This question was a single-answer question with five response choices including being infected by COVID-19, online distance learning, the economic impacts of the COVID-19 pandemic, curfew and social isolation, and other concerns.

The questionnaire was piloted on 10 students who were approached by the first author to test the phrasing, suitability, and understandability of the questions. The responses from these 10 students, as well as incomplete questionnaires, were excluded from the analysis.

### 2.3 Data Management and Analysis

Completed questionnaires were extracted from Google Forms as an Excel sheet and were then incorporated into STATA IC 16.1 (StataCorp LLC., Texas, USA). Descriptive analysis and summary statistics were used in which numerical variables were described as mean and standard deviation, while categorical variables were described as frequency and percentage. In addition, non-parametric tests were used including Wilcoxon Rank-Sum test to compare the mean of total K10 distress scores between males and females while Spearman’s rank correlation to test the relationship between age and total K10 distress scores. Besides, ordinal logistic regression was employed to assess the correlation between psychological distress severity (outcome variable with ordinal responses) and other independent sociodemographic predictors. The confidence level was set at 95% and a *P-*value less than 0.05 was considered statistically significant.

### 2.4 Ethical considerations

The study was conducted according to the *Declaration of Helsinki*. Ethical approval was granted by the Institutional Review Board at Al-Zaytoonah University of Jordan. Besides, the questionnaire ensured the privacy and confidentiality of participants by not asking any questions about names, phone numbers, physical addresses, or emails; thus, all participants were anonymous. Also, an information letter was incorporated into the first page of the questionnaire and included explicit information about the researchers and their affiliations, the study description and objectives, eligibility criteria for participation, voluntary participation and withdrawal, benefits and risks, privacy and confidentiality aspects, data handling, as well as the contact details for any enquiry. Furthermore, at the end of the information letter, electronic informed consent was requested from participants as a prerequisite to join the survey voluntarily.

## 3 RESULTS

### 3.1 Respondents’ Characteristics

A total of 397 questionnaires were received, and 16 were excluded due to incompleteness. So, the remaining 381 were included in our analysis. There was a slight predomination of female participants (n=199, 52.2%) compared to male participants (n=182, 47.8%). The mean age was 22.6 years (SD=3.16) and ranged between 18-38 years. The vast majority of participants were single (n=352, 92.4%), undergraduates (n=323, 84.8%), studying at governmental/public universities or colleges (n=209, 54.9%), living in the central region of Jordan (n=302, 79.3%), currently non-smokers (n=267, 70.1%) as well as with no history of pre-existing psychological or mental illness (n=366, 96.1%). More details about the sociodemographic characteristics of the respondents are provided in **Table 1**.

**Table 1.** Sociodemographic characteristics of the Respondents (n=381)

| Variables | Results |
| --- | --- |
| **Sex** |  |
| Male | n=182 (47.8%) |
| Female | n=199 (52.2%) |
| **Age (Mean, SD)** | **22.6 , 3.16** |
| 18-22 | n= 208 (54.6%) |
| 23-27 | n=142 (37.3%) |
| 28-32 | n=22 (5.8%) |
| 33-38 | n=9 (2.3%) |
| **Marital Status** |  |
| Single | n=352 (92.4%) |
| Married | n=29 (7.6%) |
| **Region of residence** |  |
| Northern governorates | n=60 (15.7%) |
| Central governorates | n=302 (79.3%) |
| Southern governorates | n=19 (5.0%) |
| **Smoking Status** |  |
| Current Smoker | n=117 (29.9%) |
| Currently non-smoker | n=267 (70.1%) |
| **Academic Institution** |  |
| Public university/college | n=209 (54.9%) |
| Private university/college | n= 172 (45.1%) |
| **Study Level** |  |
| Undergraduate | n=323 (84.8%) |
| Postgraduate | n=58 (15.2%) |
| **History of pre-existing psychiatric conditions** |  |
| Yes | n=15 (3.9%) |
| No | n=366 (96.1%) |
| **Current use of medications among the 15 students who reported a history of pre-existing psychiatric conditions** |  |
| Yes | n=8 |
| No | n=7 |

### 3.2 Kessler Psychological Distress Scale (K10) Results

The total K10 distress scores had a mean of 34.2 (SD=9.4). The mean K10 distress score was slightly higher among women (mean=34.7, SD=8.56) compared to men (mean=33.7, SD=10.3); however, Wilcoxon Rank-Sum test showed that this difference is statistically insignificant (*P*=0.566). Concerning age, Spearman’s rank correlation test revealed a statistically significant inverse relationship between age and total K10 distress score (Rho= −0.1645, *P*=0.001), which indicates that younger age groups were more likely to have higher total K10 distress scores; thus, more distress. **(Figure 1)**.

**Figure 1.**
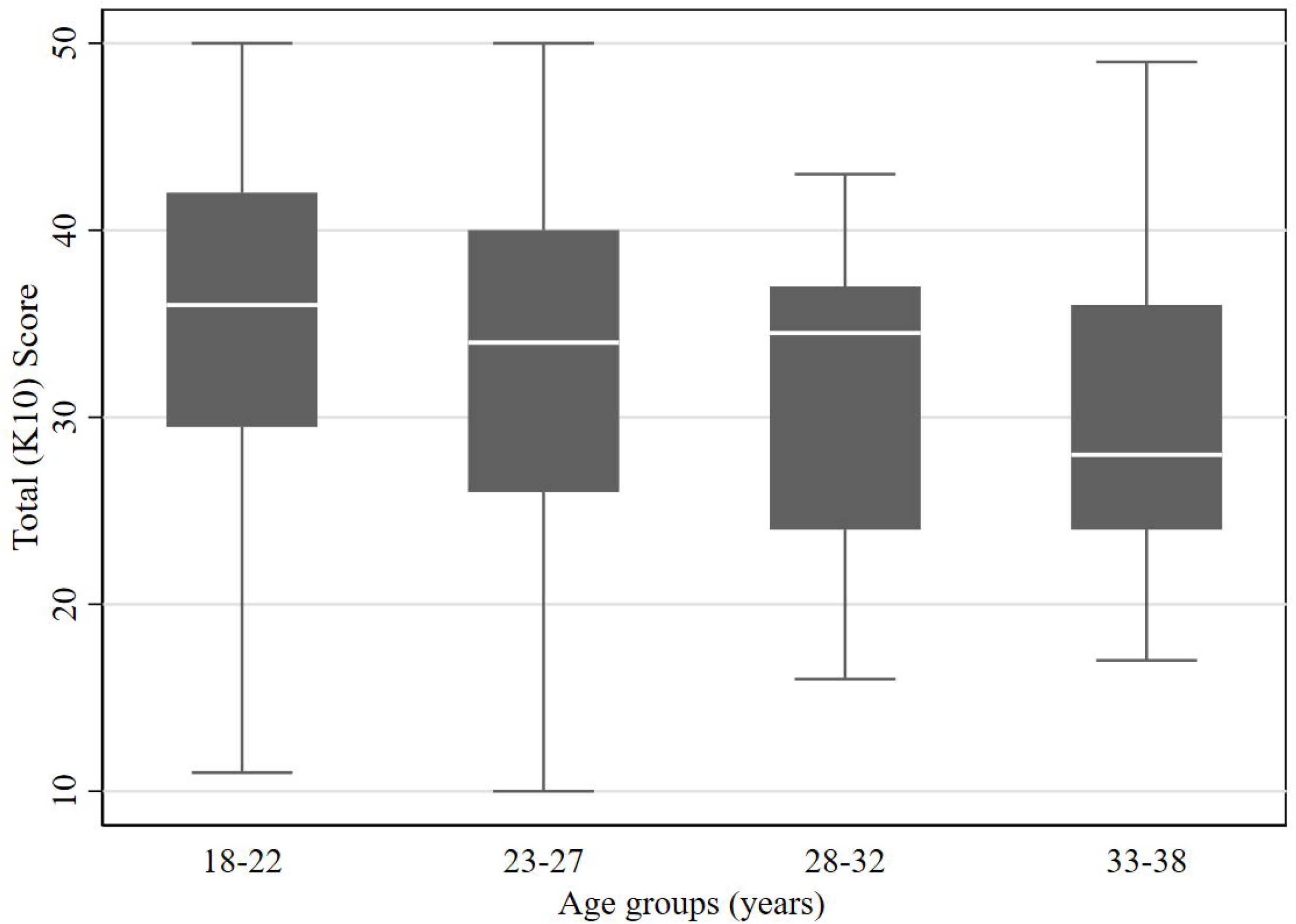
The levels of total K10 distress scores among different age groups of the respondents.

Regarding psychological distress severity categorization and based on K10 scale’s categories described earlier in study instrument, most of respondents were regarded as having severe psychological distress (n=265, 69.5%), followed by moderate psychological distress (n=48, 12.6%), mild psychological distress (n=41, 10.8%), and no psychological distress (n=27, 7.1%). **Tables 2** and **Figure 2** show more descriptive results of the K10 distress scale by severity level and gender.

**Table 2.** The severity of Psychological distress among respondents based on K10 distress scale’s categorization.

| **K10 Psychological Distress Category** | **Total K10 Score range** | **Frequency (n)** | **Percentage (%)** |
| --- | --- | --- | --- |
| **No Distress** | **10-19** | 27 | 7.1 |
| **Mild Distress** | **20-24** | 41 | 10.8 |
| **Moderate Distress** | **25-29** | 48 | 12.6 |
| **Severe Distress** | **30-50** | 265 | 69.5 |
| **Total** |  | **381** | **100** |

**Figure 2.**
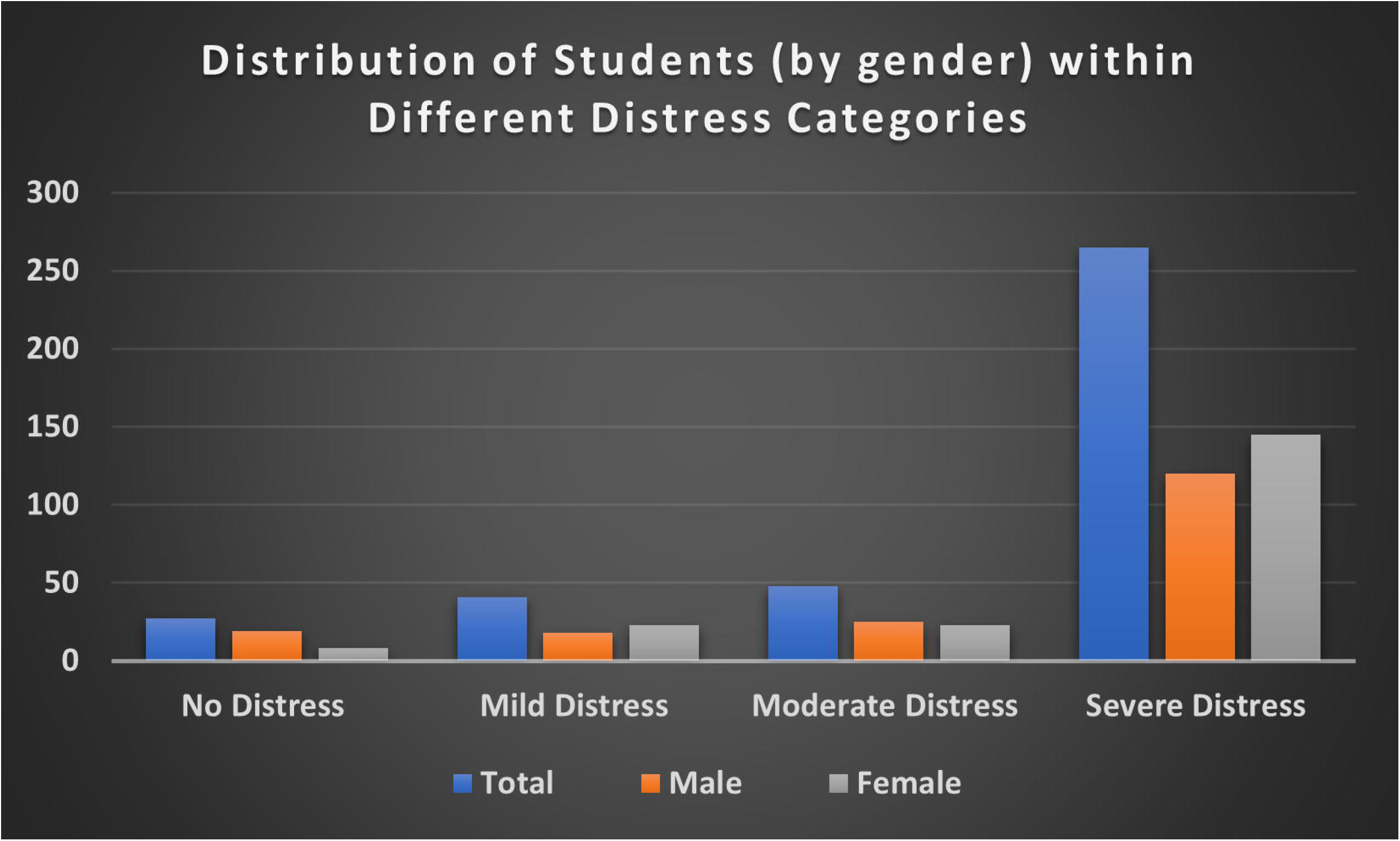
Distribution of students (by gender) within different distress categories based on the overall K10 score for each student.

Ordinal logistic regression was employed to assess the correlation between distress severity (ordinal outcome variable) and other sociodemographic predictors, however, considering our sample size (n=381) and in order to achieve sufficient statistical power for the regression test, we have merged *no distress* and *mild distress* in one ordinal category as well as *moderate distress* and *severe distress* together in another ordinal category. Therefore, we had an ordinal outcome variable with two severity levels/categories. After that, we tested each independent variable against the outcome variable. All independent variables that had a P value less than 0.25 in univariable regression were included in the final ordinal logistic regression model. The regression model revealed a significant correlation between distress severity and some predictors. Among the predictors that were found to act as a protective factor against higher levels of distress were *older age* (aOR=0.64, P=0.022; 95% CI: 0.44 – 0.94), and having a *strong motivation for distance learning* (aOR=0.10, P=0.048 ; 95% CI: 0.01 - 0.96). In contrary, being *a current smoker* (aOR=1.99, P=0.049 ; 95% CI: 1.10 - 3.39), and having *no motivation for distance learning* (aOR=2.49, P=0.007; 95% CI: 1.29 - 4.80) acted as risk factors for having higher levels of psychological distress among the students The detailed results of oridinal logistic regression are presented in **Table 3**

**Table 3.** Results of Ordinal Logistic Regression for the correlation between psychological distress severity and independent predictors.

| Predictors | Crude OR [95% CI] | P-value | Adjusted OR [95% CI] | P-value |
| --- | --- | --- | --- | --- |
| **Age** | 0.67 [0.50 - 0.90] | 0.008 | 0.64 [0.44 - 0.94] | **0.022** |
| **Gender** |  |  |  |  |
| Female | **Reference** |  | **Reference** |  |
| Male | 0.67 [0.40 - 1.1] | 0.141 | 0.56 [0.30 - 1.03] | 0.063 |
| **Smoking** |  |  |  |  |
| No | **Reference** |  | **Reference** |  |
| Yes | 1.48 [0.81 - 2.72] | 0.206 | 1.99 [1.10 - 3.39] | **0.049** |
| **Study Level** |  |  |  |  |
| Postgraduate | **Reference** |  | **Reference** |  |
| Undergraduate | 0.59 [0.26 - 1.36] | 0.216 | 0.53 [0.17 - 1.64] | 0.272 |
| **University/College** |  |  |  |  |
| Private | **Reference** |  | **Reference** |  |
| Public | 1.96 [1.15 - 3.34] | 0.013 | 1.43 [0.74 - 2.77] | 0.287 |
| **Motivation for Distance learning** |  |  |  |  |
| Low | **Reference** |  | **Reference** |  |
| No | 2.62 [ 1.40 - 4.93] | 0.003 | 2.49 [1.29 - 4.80] | **0.007** |
| Moderate | 0.99 [0.49 - 2.02] | 0.983 | 1.27 [0.59 - 2.73] | 0.535 |
| High/Strong | 0.08 [0.01 - 0.76] | 0.028 | 0.10 [0.01 - 0.98] | **0.048** |

### 3.3 The motivation for Distance Learning

Surprisingly, a significant proportion of the students have reported that they had *no motivation at all* toward the online distance learning (n=209, 54.9%),, and as described earlier, students with no motivation for distance learning were more likely to suffer from higher degrees of psychological distress (aOR=2.49, P=0.007; 95% CI: 1.29 - 4.80).**Table 4 and Figure 3** demonstrate more descriptive details about the motivation for distance learning.

**Table 4.** The degree of motivation for online distance learning among respondents.

| **Degree of Motivation** | **Frequency (N)** | **Percentage (%)** |
| --- | --- | --- |
| No Motivation | 209 | 54.9 |
| Low Motivation | 98 | 25.7 |
| Moderate Motivation | 69 | 18.1 |
| Strong Motivation | 5 | 1.3 |
| **Total** | **381** | **100** |

**Figure 3.**
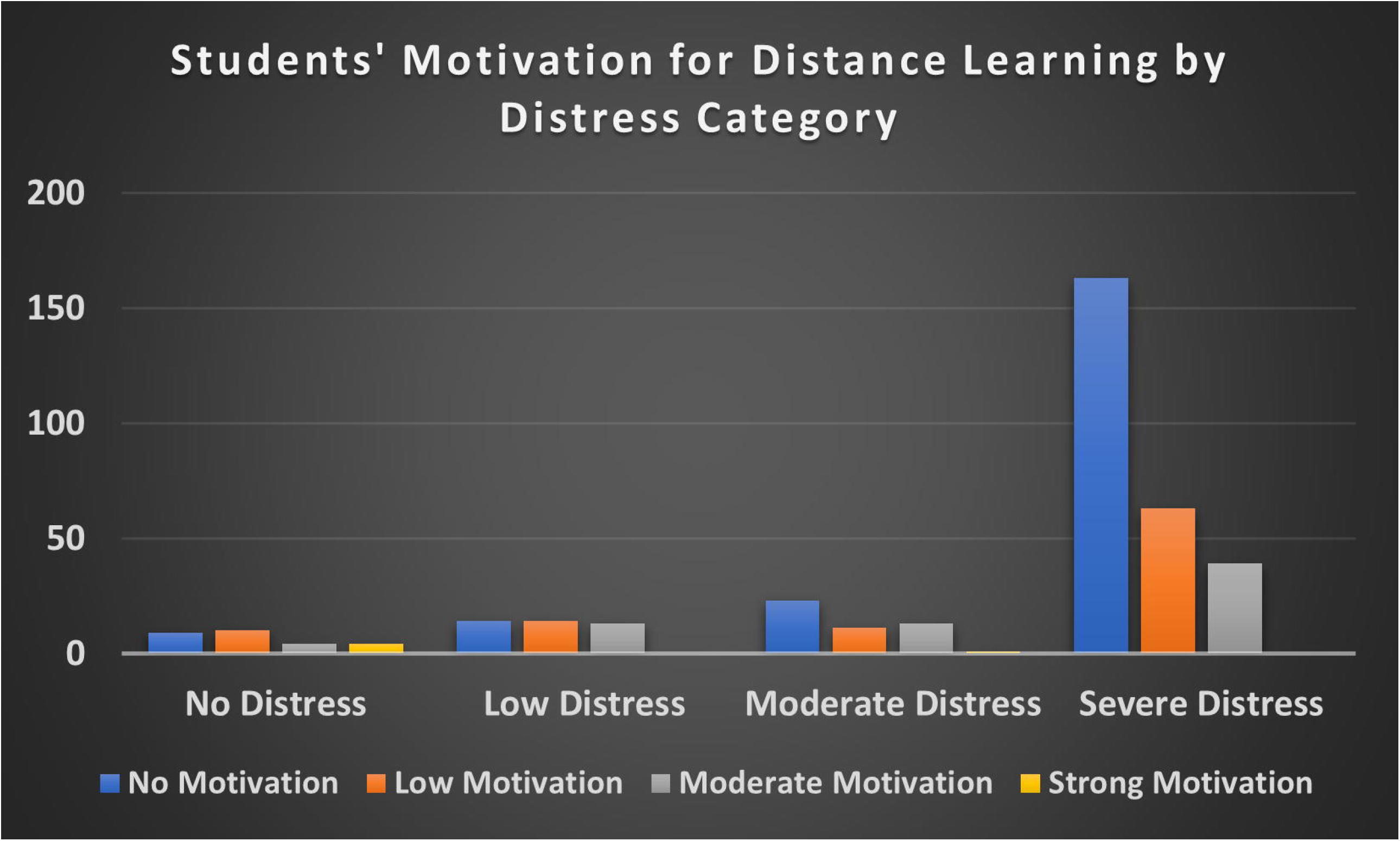
Students’ Motivation for Distance Learning per Distress Category

### 3.4 Coping Activities and Concerns during the COVID-19 Pandemic

The students have selected many coping activities that the they frequently practiced during the nationwide curfew in Jordan. Interestingly, the responses with highest frequencies were *spending more time on social networking platforms like Facebook and Instagram* (n=269, 70.6%), *talking to friends on mobile phones and internet* (n=217, 57%), *watching television and movies* (n=210, 55.1%), *more engagement with family* (n=202, 53%), and *listening to music* (n=162, 42.5%). More details about these activities are provided in **Table 5**.

**Table 5.** Coping activities during the COVID-19 pandemic and the nationwide curfew in Jordan among the respondents.

| **Coping Activity** | **Frequency (n)** | **Percentage (%)** |
| --- | --- | --- |
| Spending more time on social networking platforms like Facebook and Instagram | 269 | 70.6 |
| Talking to friends on mobile phones and internet | 217 | 57 |
| Watching television and movies | 210 | 55.1 |
| More engagement with the family | 202 | 53 |
| Listening to music | 162 | 42.5 |
| Practicing sports at home | 113 | 29.7 |
| Studying and preparing for exams | 102 | 26.8 |
| Increase smoking | 69 | 18.1 |
| Reading Books / Novels | 68 | 17.8 |
| Meditation | 58 | 15.2 |
| Herbal drinks | 57 | 15 |
| Practicing Yoga | 6 | 1.6 |
| Talking to a psychological counselor | 6 | 1.6 |
| Others | 33 | 8.7 |

In addition, among the 381 respondents, 332 students (87.1%) reported no use of any medications during the last 30 days for coping with the distress accompanied the COVID-19 pandemic and the nationwide curfew, while 49 students (12.9%) reported the use of various types of medications at different frequencies with occasionally (1-2 times in a month) as the most common frequency. Sedative hypnotics (38%) reported being on the top of the used medications followed by others (28%), which included over-the-counter medications like Paracetamol and other simple analgesics. More details are demonstrated in **Figures 4, 5**, and **Table 6**.

**Table 6.** Medicinal drugs’ usage frequency among the 49 students who reported the use of different medications in response to the COVID-19 induced distress.

| **Frequency of usage** | **Number of students** | **Percentage (%)** |
| --- | --- | --- |
| 1-2 times in a month | 17 | 34.7 |
| 1-2 times in a week | 13 | 26.5 |
| 3-4 times in a week | 10 | 20.4 |
| Everyday | 9 | 18.4 |
| **Total** | **49** | **100** |

**Figure 4.**
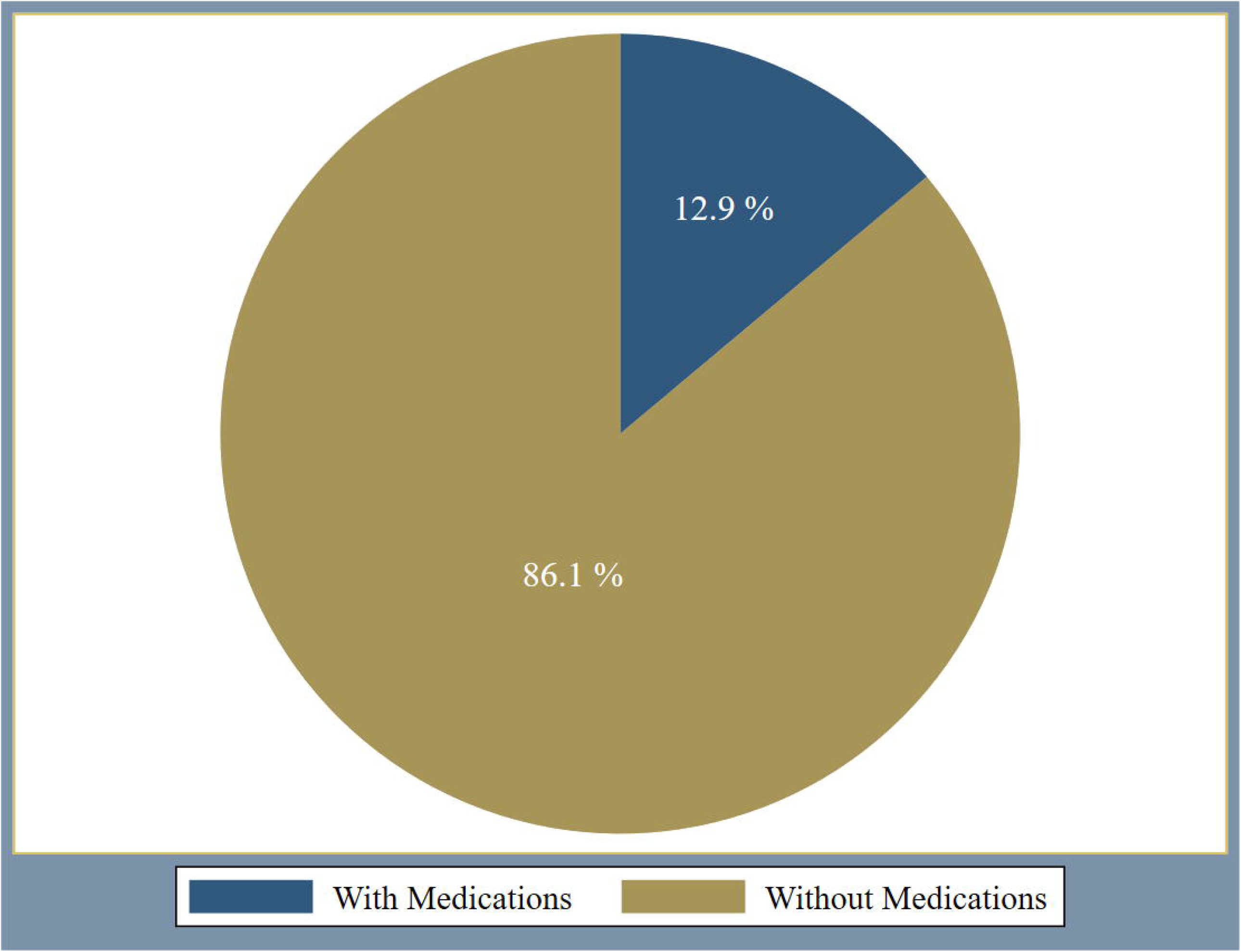
Reported medication use for coping with the COVID-19 related psychological distress among respondents (percentage)

**Figure 5.**
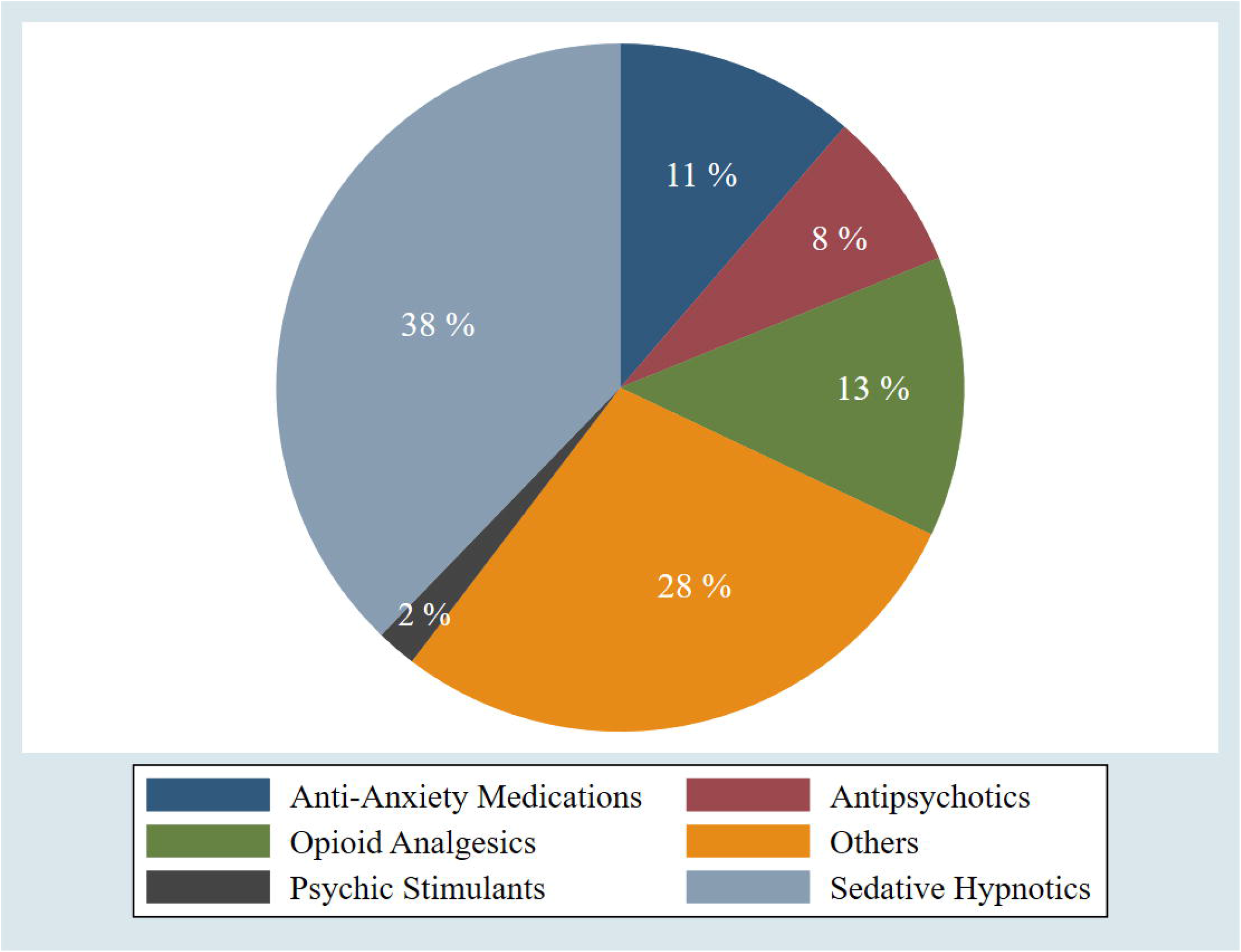
Types of medications that were used by 49 students for coping with the COVID-19 induced psychological distress.

Moreover, 209 students (54.9%) reported that *online distance learning* was the highest and most serious issue of concern, followed by 75 students (19.7%) who reported *curfew and social isolation* as their highest issue of concern. Unexpectedly, only 53 students (13.9%) reported *being infected by COVID-19* as their most serious concern. **Figure 6** for more illustration.

**Figure 6.**
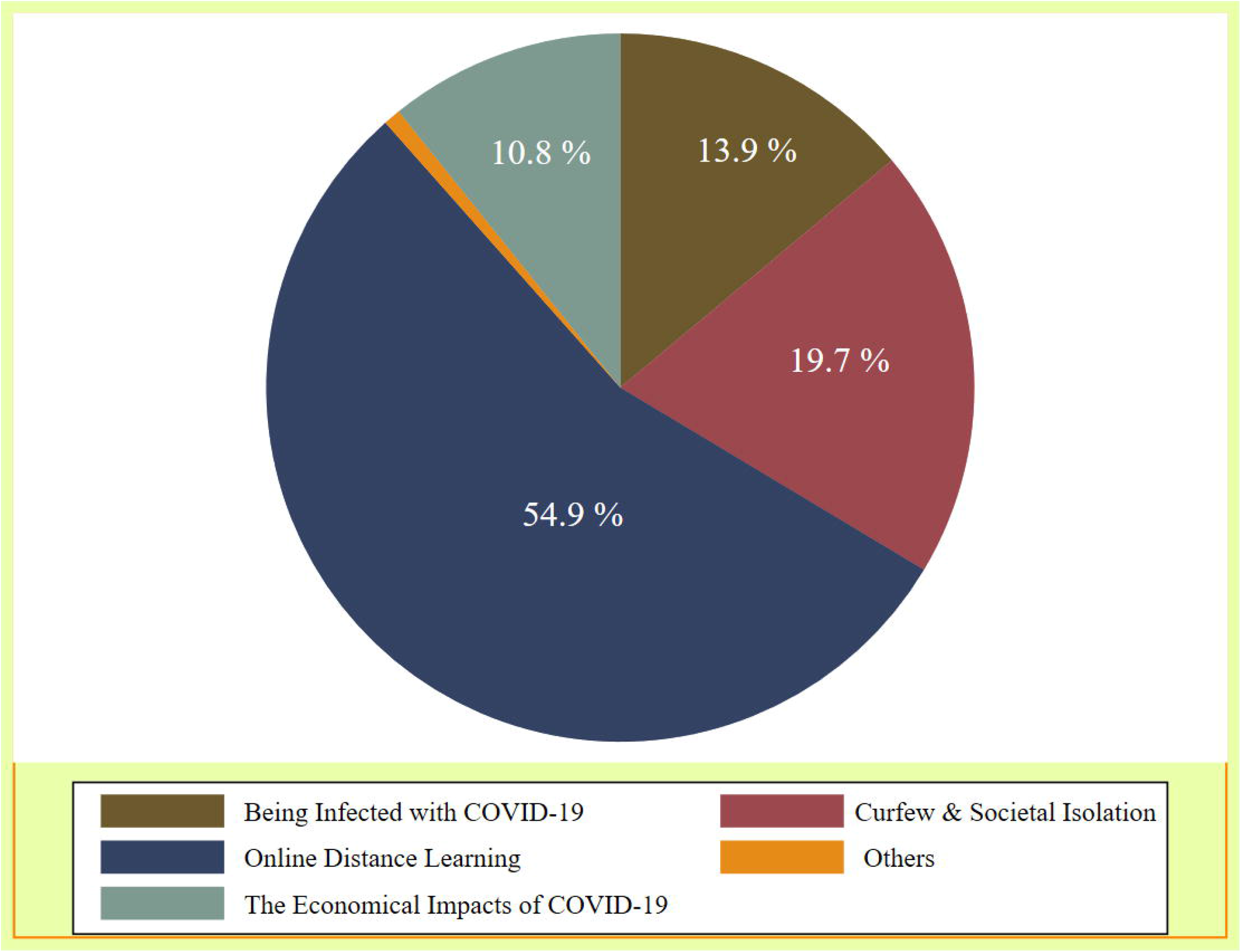
The issue of the greatest concern as perceived by the 381 students (percentage)

## 4 DISCUSSION

In our study, the vast majority of the students (92.9%) suffered from different levels of psychological distress ranging from mild to severe degrees during the COVID-19 pandemic.The psychological wellbeing of university students in the midst of the current pandemic has been established and reported in recently published litratures as well. A recent study which was conducted by Cao et al. (2020) in China and aimed at exploring the psychological impact of COVID-19 on college students using the 7-item Generalized Anxiety Disorder Scale (GAD-7) has revealed that 24.9% of students suffered from anxiety during this pandemic with a positive association of the level of anxiety with different economic and academic stressors (Cao et al., 2020). Similar to Cao et al. (2020) study, our study found that there was no significant difference in the total psychological distress scores between men and women. In addition, Cao et al. found that social support was negatively associated with anxiety status among students, and we have that as one of our most reported coping mechanisms, i.e., socialization through social networking sites. Nevertheless, in our study, age was statistically and significantly associated with distress severity; i.e the younger the age, the more likely to suffer from higher levels of psychological distress. The difference in distress proportions between our study (92.9%) and Cao et al study (24.9 %) could be attributed to the use of different scales, i.e. GAD-7 vs. K10 as well as the sample size. In addition, we carried out the survey in a period close to final examinations, which might have had an additional negative impact on the students’ psychological status.

Additionally, a recent study by Olimat et al (2020) (Olaimat et al., 2020) was conducted to assess attitudes, anxiety, as well as behavioral practices among university students in Jordan amidst the COVID-19 pandemic using an online survey developed by the authors to serve their study objectives. The study has found that 69.2 % of participants reported being anxious as a result of fear of infection by COVID-19 and resultant disruptions in their lives.Among the predicitve factors that affected the students’ anxiety levels were age, gender and academic discipline of their study programs.Older students and female students were found to have more anxiety due to the fear of infection. However, in our present study the mean total K10 distress score was higher among women compared to men, but this difference in means was statistically not significant. In contrary, older age was amongst the protective factors against higher levels of psychological distress in our present study.

Moreover, a cross sectional study was conducted in Turkey which aimed at assessing anxiety status of university students using an online survey.The measure of anxiety levels were obtained using the Turkish version of abbreviated Beck Anxiety Inventory.The study has found that 44 % of students reported a moderate level of worrisome and fear of catching COVID-19, while 80% of students reported a ‘severe level’ of scare and worries about thier close relatives’ health. The authors expected that the high levels of anxiety among the students in their study could be attributed to shifting to online learning alongwith other pandemic control measures such as social isolation and financial constraints (Akdeniz et al., 2020). Similarly, the aforementioned worries were also reported by the students in our study as part of their major concerns during the COVID-19 pandemic.

Furthermore, Stress and anxiety were assessed in France among university students during the current pandemic.University students were asked to complete the World Mental Health International College Student survey which was distributed as an online survey.Among the 291 participants in the study, the majority of them experienced significant proportions of psychological distress of which 60.2% of students reported escalation of their anxiety to moderate-severe levels during the COVID-19 pandemic.However, 82.2% of the students in our study reported moderate to severe distress. This difference in distress protportions coud be as a result of different scales used and cross-cultural factors.In the same study, the researchers found significant factors that affected the students’ anxiety level including the economic situation of the students, and the disruptions in students’ life (Husky et al., 2020). These factors were also reported in our study as pandemic induced concerns as perceived by the students. However, in our study, we have not collected data about the students’ or their families financial status ; thus we could not consider it in our regression analysis to examine its influence on the distress levels.

Jordanian universities have been taking humble attempts to implement distance learning into their educational system since 2015. Nevertheless, this strategy has been considered as a “challenging pedagogy” of the learning system in the country due to many obstacles (Al-Jaghoub et al., 2009; Atoum et al., 2017; Al Nawas, 2020). During the COVID-19 pandemic and after realising the need to implement an emergency distance learning strategy, more serious steps were taken by decision makers at higher education sector and the Jordanian universities trying to guarantee a smooth shifting process coupled with ensuring a quality education as well. Besides, psychological distress was reported to be associated with distance learning and working from homes during the current pandemic. A recent qualitative study has addressed many of the distance learning’s challenges including personal, technological, course-related as well as cultural challenges (Almaiah et al., 2020).These challenges might explain why most of the students (n=209, 45.9%) in our study resported the lack of motivation for distance learning, especially within the Jordanian context where most of educational activities were used to be delivered by in-person attendance to universities/colleges with less attention to distance learning.

Besides, smoking exhibited a risk factor for suffering from higher levels of psychological distress among the students in our study, and this could be explained by the *bi-directional* relatiohship between smoking and mental wellbeing as addressed previously in a longitudinal study in Australia (Leung et al., 2012). Emotional and behavioural reactions toward the COVID-19 pandemic could vary. The type of coping strategy and the extent of adopting it also differs between individuals. In the present study, some students (12.9%) reported the use of various medicinal drugs as a result of pandemic induced distress. Although the figure is small but this raises a concern about the psychosocial response of some individuals in response to crisis which might lead to a risky behaviour such as substance abuse.Therefore, more serious efforts should be done to spread awareness about healthy coping styles among different social components of the community (Pfefferbaum and North, 2020).

To the best of our knowledge, this is the first study in Jordan to assess the psychological distress among university students using the 10-item Kessler Psychological Distress Scale (K10) during the COVID-19 pandemic. In addition, this study is amongst the limited litratures to highlight the distressing concerns brought about by online distance learning on university students in Jordan. Still, there are limitations that should be carefully taken into consideration when interpreting the results and include (i) using a non-probabilistic convenience sampling, which affects the representativness of our sample and limits the generalizability of our results. However, this sampling strategy was believed to fit in lieu of the current circumstances of the nationwide curfew, the closure of all universities and colleges in the country and shifting to online platforms (ii) the majority of respondents were undergraduates; we could have seen different results if our sample had more postgraduate students, and (iii) We had a relatively small sample size which could be attributed to the limited period of data collection. There was a technical difficulty to follow up the survey and keep it visible to students within social media groups due to the large number of academic enquiries posted on these groups; thus, enforced our survey link to lose its visibility among the numerous recent posts. Also, the busy schedule of students (in the midst of a new distance learning strategy) might have affected their interests to participate in the survey (iv) The survey represented self reported states thus, over reporting or underreporting of psychological status could be expected, (v) The inherent limitation of cross-sectional studies which prevent assessing temporality of events i.e psychogoligcal distress could be present prior to the pandemic and just escalated during it, and lastly, (vi) We missed the perspectives of non-arabic speaking students in Jordan as the questionnaire was designed in Arabic only. Nevertheless, findings from our study shed the lights on various degrees of psychological distress that the university students have experienced during the current pandemic, and they could be considered as a vulnerable group. Also, the findings of our study encourage for further follow up research on this topic using a nationally representative sample of university students with more specific scales for psychological distress symptoms.

The results of this study provide new insights to direct policy makers and decision makers in the fields of higher education, as well as mental health. More attention and monitoring of college students’ mental health should be sought. Since distance learning was the highest reported concern among students, faculty members should implement effective methods to make distance learning more interactive and students friendly. Psychological interventions should be implemented by psychologists and psychiatrists to provide guidance, psychoeducation, and mental health counseling to university students. There should be more active involvement with students’ psychological health, coupled with educating them on how to deal with psychological distress during unprecedented situations like the current pandemic.

At the current circumstances of COVID-19 preventive measures in Jordan (curfew and social distancing), psychological support could be provided to university students through publicly available online videos, television programs, and online/phone consultations. Also, mental health support could be provided through a hotline service to provide students with instructions about dealing with their academic stressors and other related mental health issues during this pandemic.

Moreover, efforts should be made to improve communications with college students’ and guide them on how to access only evidence-based information from reliable resources about the pandemic. Besides, a comprehensive nationwide psychological support program should be developed and incorporated into Jordan’s response strategy in combating the COVID-19. Future studies should assess the effect of implementing these suggested interventions on students’ mental health. Furthermore, as the levels of psychological distress are expected to be dynamic over the upcoming period, it is wise to monitor and assess the impact of easing up the governmental restrictions, i.e. ending the curfew and returning to on-campus teaching, on the levels of psychological distress and anxiety among university students in Jordan.

## 5 CONCLUSION

The control and preventive measures that are implemented during the COVID-19 pandemic resulted in a severe disruption of various human life activities. The fear of the infection itself, along with the strict public health measures could impact the mental health of individuals. Our study highlighted a significant psychological distress among university students in Jordan during the COVID-19 pandemic and its related control measures.. A significant proportion of the students were highly concerned about and distressed by the distance learning strategy; thus, prompt actions should be taken to improve the distance learning experience and solve any associated technostress. In addition, a nationwide psychological support program should be incorporated into Jordan’s preparedness plan and response strategy in combating the COVID-19 pandemic and other crisis, considering students and other vulnerable groups in the community.

## Data Availability

The dataset generated and analyzed for this study are available from the corresponding author on a reasonable request

## Acknowledgments

The authors provide their sincere appreciation to university students who participated in the online survey. Also, Al-Tammemi gratefully acknowledges the funding received from The Swedish Institute toward his study at the Department of Epidemiology and Global Health, Umeå University as well as the funding received from Tempus Public Foundation toward his doctoral study at the Doctoral School of Health Sciences, University of Debrecen. The funders had no role in the study design, data collection and analysis, decision to publish, or preparation of the manuscript.

## Authorship statement

Al-Tammemi conceptualized the study, designed, and prepared the questionnaire with inputs from Akour and Alfalah. Data collection was carried out by Al-Tammemi and Akour. The statistical analyses were done by Al-Tammemi.

Alfalah wrote the introduction. Al-Tammemi wrote the methods, materials, and results. Al-Tammemi and Akour wrote the discussion. All authors have substantially and critically contributed to editing and revising the manuscript and providing critical feedback. All authors have approved the submission of this version of the manuscript.

## References

Akdeniz, G., Kavakci, M., Gozugok, M., Yalcinkaya, S., Kucukay, A., and Sahutogullari, B. (2020). A Survey of Attitudes, Anxiety Status, and Protective Behaviors of the University Students During the COVID-19 Outbreak in Turkey. Front. Psychiatry 11, 695. doi:10.3389/fpsyt.2020.00695.

Al-Jaghoub, S., Al-Yaseen, H., Hourani, M., and El-Haddadeh, R. (2009). E-learning adoption in higher education in Jordan: vision, reality and change. in European and Mediterranean Conference on Information Systems - EMCIS 2009 Available at: https://bura.brunel.ac.uk/bitstream/2438/4044/1/plugin-C83.pdf.

Al-Tammemi, A. B. (2020). The Battle Against COVID-19 in Jordan : An Early Overview of the Jordanian Experience. Front. Public Heal. 8, 188. doi:10.3389/fpubh.2020.00188.

Al Nawas, B. (2020). Higher Education Council reviews distance learning experience. The Jordan Times. Available at: http://jordantimes.com/news/local/higher-education-council-reviews-distance-learning-experience.

Almaiah, M. A., Al-Khasawneh, A., and Althunibat, A. (2020). Exploring the critical challenges and factors influencing the E-learning system usage during COVID-19 pandemic. Educ. Inf. Technol. doi:10.1007/s10639-020-10219-y.

Andrews, G., and Slade, T. (2001). Interpreting scores on the Kessler Psychological Distress Scale (K10). Aust. N. Z. J. Public Health 25, 494–497. doi:10.1111/j.1467-842X.2001.tb00310.x.

Atoum, A., Al-Zoubi, A., Jaber, M. A., Al-Dmour, M., and Hammad, B. (2017). A New Approach for Delivering eLearning Courses in Jordanian Universities. Adv. Soc. Sci. Res. J. 4, 1–13. doi:10.14738/assrj.48.3071.

Brooks, S. K., Webster, R. K., Smith, L. E., Woodland, L., Wessely, S., Greenberg, N., et al. (2020). Rapid Review The psychological impact of quarantine and how to reduce it: rapid review of the evidence. Lancet 395, 912–920. doi:10.1016/S0140-6736(20)30460-8.

Cao, W., Fang, Z., Hou, G., Han, M., Xu, X., Dong, J., et al. (2020). The psychological impact of the COVID-19 epidemic on college students in China. Psychiatry Res. 287. doi:10.1016/j.psychres.2020.112934.

Center for the Study of Traumatic Stress (2020). Psychological Effects of Quarantine During the Coronavirus Outbreak: What Healthcare Providers Need to Know. Available at: https://www.cstsonline.org/assets/media/documents/CSTS_FS_Psychological_Effects_Quarantine_During_Coronavirus_Outbreak_Providers.pdf [Accessed May 5, 2020].

Chakraborty, I., and Maity, P. (2020). COVID-19 outbreak: Migration, effects on society, global environment and prevention. Sci. Total Environ. 728, 138882. doi:10.1016/j.scitotenv.2020.138882.

Easton, S. D., Safadi, N. S., Wang, Y., and Hasson, R. G. (2017). The Kessler psychological distress scale: Translation and validation of an Arabic version. Health Qual. Life Outcomes 15, 215. doi:10.1186/s12955-017-0783-9.

Fassaert, T. J. L., Wit, M. A. S. De, Tuinebreijer, W. C., Wouters, H., Verhoeff, A. P., Beekman, A. T. F., et al. (2009). Psychometric Properties of an interviewer-administered version of the Kessler Psychological distress scale (K10) among dutch, moroccan and turkish respondents. Int. J. Methods Psychiatr. Res. 18, 159–168. doi:10.1002/mpr.288.

Ho, C. S., Chee, C. Y., and Ho, R. C. (2020). Mental Health Strategies to Combat the Psychological Impact of COVID-19 Beyond Paranoia and Panic. Ann. Acad. Med. Singapore 49, 1–3.

Holmes, E. A., O’connor, R. C., Perry, H., Tracey, I., Wessely, S., Arseneault, L., et al. (2020). Position Paper Multidisciplinary research priorities for the COVID-19 pandemic: a call for action for mental health science. Lancet Psychiatry. doi:10.1016/S2215-0366(20)30168-1.

Husky, M. M., Kovess-Masfety, V., and Swendsen, J. D. (2020). Stress and anxiety among university students in France during Covid-19 mandatory confinement. Compr. Psychiatry 102, 152191. doi:10.1016/j.comppsych.2020.152191.

Jordanian Ministry of Health (2020). COVID-19 in Jordan. [Online]. Available at: https://corona.moh.gov.jo/ar [Accessed April 16, 2020].

Kessler, R. C., Andrews, G., Colpe, L. J., Hiripi, E., Mroczek, D. K., Normand, S. L. T., et al. (2002). Short screening scales to monitor population prevalences and trends in non-specific psychological distress. Psychol. Med. 32, 959–976. doi:10.1017/S0033291702006074.

Leung, J., Gartner, C., Hall, W., Lucke, J., and Dobson, A. (2012). A longitudinal study of the bi-directional relationship between tobacco smoking and psychological distress in a community sample of young Australian women. Psychol. Med. 42, 1273–1282. doi:DOI: 10.1017/S0033291711002261.

National Comorbidity Survey (2013). National Comorbidity Survey: Arabic K10. [Online]. Available at: https://www.hcp.med.harvard.edu/ncs/k6_scales.php [Accessed May 2, 2020].

Olaimat, A. N., Aolymat, I., Elsahoryi, N., Shahbaz, H. M., and Holley, R. A. (2020). Attitudes, Anxiety, and Behavioral Practices Regarding COVID-19 among University Students in Jordan: A Cross-Sectional Study. Am. J. Trop. Med. Hyg. 103, 1177–1183. doi:10.4269/ajtmh.20-0418.

Ornell, F., Schuch, J. B., Sordi, A. O., and Kessler, F. H. P. (2020). “Pandemic fear” and COVID-19: mental health burden and strategies. Brazilian J. Psychiatry 00, 000–000. doi:10.1590/1516-4446-2020-0008.

Pfefferbaum, B., and North, C. S. (2020). Mental Health and the Covid-19 Pandemic. N. Engl. J. Med. 383, 510–512. doi:10.1056/NEJMp2008017.

Prime Ministry of Jordan (2020). Official Reports. [Online]. Available at: http://www.pm.gov.jo/category/7603/اخبار.html [Accessed April 17, 2020].

SAMHSA (2014). Coping With Stress During Infectious Disease Outbreaks. Available at: https://store.samhsa.gov/sites/default/files/d7/priv/sma14-4885.pdf [Accessed May 5, 2020].

Thornton, L., Batterham, P. J., Fassnacht, D. B., Kay-Lambkin, F., Calear, A. L., and Hunt, S. (2016). Recruiting for health, medical or psychosocial research using Facebook: Systematic review. Internet Interv. 4, 72–81. doi:10.1016/j.invent.2016.02.001.

USCF (2020). Emotional Well-Being and Coping During COVID-19. [Online]. Available at: https://psychiatry.ucsf.edu/copingresources/covid19.

World Health Organization (2020). Coronavirus disease (COVID-19) outbreak situation. [Online]. Available at: https://www.who.int/emergencies/diseases/novel-coronavirus-2019 [Accessed June 25, 2020].

